# Statistical learning in patients in the minimally conscious state

**DOI:** 10.1101/2022.01.04.22268656

**Authors:** Chuan Xu, Jian Gao, Jiaxin Gao, Lingling Li, Fangping He, Jie Yu, Yi Ling, Hangcheng Li, Jingqi Li, Lucia Melloni, Benyan Luo, Nai Ding

**Affiliations:** Department of Neurology, First Affiliated Hospital, School of Medicine, Zhejiang University, Hangzhou 310003, China; Department of Rehabilitation, Hangzhou Mingzhou Brain Rehabilitation Hospital, Hangzhou 311215, China; Key Laboratory for Biomedical Engineering of Ministry of Education, College of Biomedical Engineering and Instrument Sciences, Zhejiang University, Hangzhou 310027, China; Research Center for Advanced Artificial Intelligence Theory, Zhejiang Lab, Hangzhou 311121, China; New York University Comprehensive Epilepsy Center, 223 34th Street, New York, NY 10016, USA

**Keywords:** statistical learning, disorders of consciousness, stimulus-locked activity

## Abstract

When listening to an unknown language, listeners could learn the transitional probability between syllables and group frequently co-occurred syllables into a whole unit. Such statistical learning ability has been demonstrated for both pre-verbal infants and adults, even during passive listening. Here, we investigated whether statistical learning occurred in patients in minimally conscious state (MCS) and patients emerged from the minimally conscious state (EMCS) using electroencephalography (EEG). We presented to participants an isochronous sequence of syllables, which were composed of either 2-word real phrases or 2-word artificial phrases that were defined by the transitional probability between words. An inter-trial phase coherence (ITPC) analysis revealed that the phrase-rate EEG response was weakened in EMCS patients compared with healthy individuals, and was even more severely weakened in MCS patients. Although weak, the phrase-rate response or its harmonics remained statistically significant in MCS patients, suggesting that the statistical learning ability was preserved in MCS patients. The word-rate response was also weakened with a decreased level of consciousness. The harmonics of the word-rate response, however, were more salient in MCS than EMCS patients in the alpha and beta bands. Together with previous studies, the current results suggest that MCS patients retain residual learning ability, which can potentially be harnessed to induce neural plasticity, and that different frequency bands are differentially related to the consciousness level.

## Introduction

The ability to learn the statistical regularities in an environment is critical for the survival of biological organisms. Statistical learning has been demonstrated for different sensory modalities, tasks, and species [1] and hypothesized as a critical building block for higher-level cognitive functions [1, 2]. In the domain of language processing, it has been established that 8-month-old infants can learn to segment a sequence of syllables into multi-syllabic words by exploiting the transitional probability between syllables [3]. Behaviorally, both infants and adults rate the multi-syllabic words respecting the transitional probabilities they are exposed to as being more familiar than other combinations of syllables that while present in the stimulus materials do not respect the transitional probabilities. It has been hypothesized that statistical learning is a fundamental mechanism for infants to discover larger linguistic units such as words and phrases from running speech.

For healthy infants and adults, statistical learning has been demonstrated during passive listening [3, 4] and can be enhanced when participants attend to the syllable sequence [5]. Nevertheless, it remains possible that participants spontaneously attend to the syllable sequence even during passive listening. Moreover, it is postulated that unlimited associative learning (UAL) provides a general marker of consciousness [6]. Therefore, here, we investigate whether statistical learning can occur in patients in a MCS state, who have minimal language ability, attention and low-level awareness. Since MCS patients can barely make behavioral responses, we objectively quantify whether they can achieve statistical learning using EEG.

When listening to a known language, cortical activity measured by EEG or MEG can track the rhythms of multiple levels of linguistic units such as syllables, words, and phrases [7, 8]. During statistical learning, cortical tracking of artificial words or phrases quickly emerge [9-11]. Since the word/phrase-tracking EEG/MEG response can be objectively measured without any behavioral response, it can be conveniently applied to study statistical learning in difficult-to-test populations. For example, a recent study [12] finds EEG evidence for statistical learning in preverbal infants. Another recent study finds that the EEG response tracking phrases is marginally significant when MCS patients listen to their native language [13]. Here, we apply the same EEG-based method to investigate whether statistical learning can occur in the MCS, comparing the EEG responses to known phrases and phrases acquired through statistical learning.

## Methods

### Participants

This study involved 19 MCS patients (14 males; 54.58 ± 11.48 years) and 22 EMCS (17 males; 53.32 ± 12.16) patients. The following inclusion criteria were used: (i) diagnosis with MCS, EMCS based on Coma Recovery Scale-Revised (CRS-R) assessments carried out by DoC experts [14]; (ii) more than 1 month since brain injury; (iii) age over 18 years; (iv) no history of hearing impairment before brain injury; (v) presence of the auditory startle reflex; (vi) no centrally acting drugs, neuromuscular function blockers, or sedation within 24 hours prior to the study; (vii) no obvious skull bone defects (CT); (viii) no history of neurodegenerative diseases such as Alzheimer’s disease and Parkinson’s disease before brain injury. The patients suffered from traumatic brain injury (TBI), anoxic brain injury, or cerebrovascular disease.

Twenty age-matched healthy individuals (3 males; 50.45 ± 8.87) with no history of neurological diseases were also included. There was no significant age difference between healthy individuals and either of the 2 patient populations (one-way ANOVA, P = 0.485). The study was approved by the Ethical Committee of the First Affiliated Hospital of Zhejiang University, and by Hangzhou Mingzhou Brain Rehabilitation Hospital. Written informed consent was provided by the participants or their legal guardians for the experiments and the publication of their individual details in this study.

### Stimuli

The stimulus consisted of an isochronous sequence of monosyllabic Chinese words. Two conditions were presented. In the real phrase condition, the monosyllabic words constructed 4 common 2-word phrases, i.e., ‘wake up’, ‘go home’, ‘eat rice’, and ‘be quick’. In the artificial phrase condition, the monosyllabic words constructed 4 artificial phrases that were reversed real phrases, i.e., ‘up wake’, ‘home go’, ‘rice eat’, ‘quick be’. None of the artificial phrases were meaningful expressions in Chinese. Monosyllabic words were individually generated using an artificial speech synthesizer and adjusted to the same intensity and the same duration, i.e., 500 ms, following the procedure described in Ding et al. (2016) [7]. In previous studies, the syllable duration was between 250 and 330 ms [13, 15], here we further slowed down the stimulus since DoC patients might not be able to follow speech at a normal rate. When constructing the sequences, the synthesized monosyllabic words were directly concatenated, without any additional pause in between. Therefore, monosyllabic words and phrases were isochronously presented at 2 Hz and 1 Hz, respectively.

### Procedures

All participants passively listened to isochronous speech sequences while their EEG responses were recorded. All participants had their eyes open at the beginning of the experiment. Healthy individuals and EMCS patients were asked to keep still during the experiment. No other tasks instructions were given. Each condition was composed of 108 trials, and each trial consisted of 11 phrases (11s in duration). All trials in each condition were presented with a randomized order. The real and artificial phrase conditions were presented successively and the order was counterbalanced across participants within each participant group.

### EEG recording and preprocessing

EEG signals were recorded using a 64-electrodes BrainCap (Brain Products DmbH, Munich, Germany), and one electrode was placed under the right eye to record the electrooculogram (EOG). EEG signals were referenced online to FCz, but were referenced offline to the average of the bilateral mastoids. The EEG signals were filtered online with a 50-Hz notch filter to remove line noise (12th order zero-phase Butterworth filter), a low-pass antialiasing filter (70-Hz cutoff, 8th order zero-phase Butterworth filter), and a high-pass filter to prevent slow drifts (0.3-Hz cutoff, 8th order zero-phase Butterworth filter). The signals were sampled at 1 kHz and down-sampled to 80 Hz. EOG artifacts were regressed out based on the least-squares method. The EEG signal was processed following the procedure in Zou et al. (2021) [16]. All preprocessing and analyses in this study were conducted in the MATLAB software (The MathWorks, Natick, MA).

### Frequency-domain analysis

Each trial was 11 s in duration but in the frequency-domain analysis the first second of recording was discarded to remove the onset response. The remaining 10-s of data were transformed into the frequency domain using discrete Fourier transform (DFT), resulting in a frequency resolution of 0.1 Hz. The ITPC was defined as:

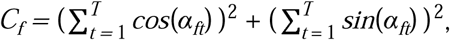

where *α*_*ft*_ denoted the DFT phase at frequency *f*, and *C*_*f*_ denoted the inter-trial phase coherence (ITPC). A higher ITPC value indicated that the response phase was consistent across trials. The ITPC at each frequency was further normalized by subtracting the mean ITPC value averaged over 2 neighboring frequency bins..

### Statistics

#### Phase coherence

This study used bias-corrected and accelerated bootstrap for all significance tests [17]. In the bootstrap procedure, all the participants were resampled with replacement 10^4^ times. The chance-level phase coherence for each target frequency was the average of the phase coherence between 0.6 and 39.5 Hz (389 frequencies were used in total, 1/10 Hz for each bin). For the significance tests for the target peaks, such as 1Hz, 2Hz, 3Hz, 4Hz….30Hz, the response amplitude at the peak frequency was compared with the mean amplitude of the 389 frequency bins. If the phase coherence at target frequency was stronger than the mean coherence of the mean coherence *N* times in the resampled data, the significance level could be calculated as (*N* + 1)/10001.

#### Bootstrap

For the unpaired comparisons between groups, we also used bias-corrected and accelerated bootstrap [17]. The data from each group were resampled with replacement 10000 times. This comparison was one-sided. If in *N* out of 10000 times, the mean in one group was greater (or smaller) than the other group, the significance level was (*N* + 1)/10001.

## Results

We analyzed the EEG responses to real and artificial phrases in 3 groups of participants, i.e., MCS patients, EMCS patients, and healthy individuals. Since the monosyllabic words were presented at an isochronous rate, the neural responses to words and the 2-word phrases were frequency tagged at 2 Hz and 1 Hz, respectively (Fig. 1). For real phrases, previous studies have shown that EEG activity can track both the rhythms of words and phrases [7, 13]. The artificial phrases were unknown to the participants at the beginning of the experiment but could be learned via statistical learning at least for healthy individuals [3] and led to a phrase-tracking neural response. We would analyze in the following whether the word and phrase responses were present in DoC patients and tested whether the strength of these responses differ across participant groups.

**Figure 1.**
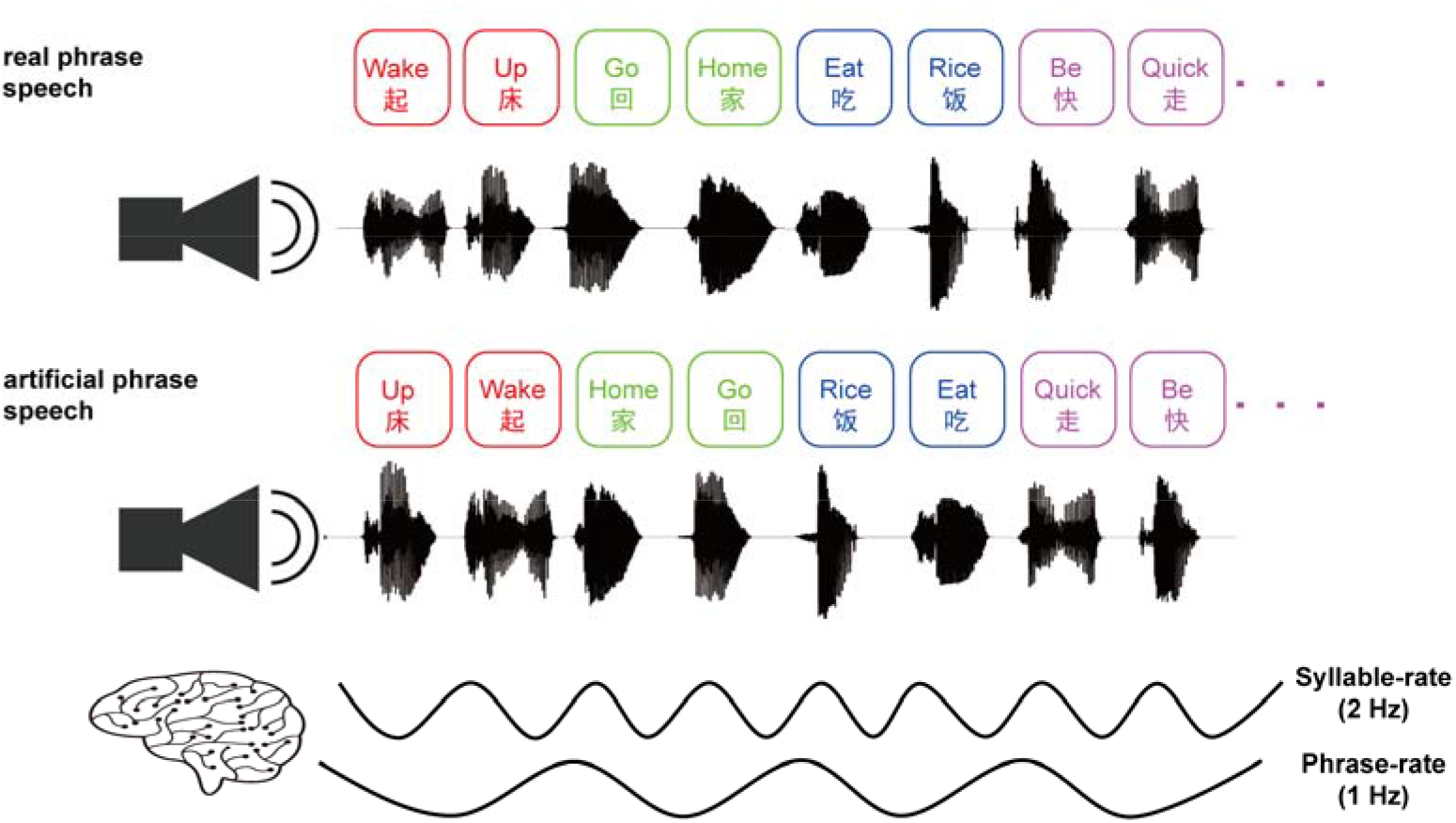
Stimulus. The monosyllabic words were presented at a constant rate of 2 Hz, corresponding to 500 ms stimuli onset asynchrony (SOA). In the real phrase condition, the stimulus was composed of 4 common 2-word phrases. In the artificial phrase condition, in contrast, the stimulus was composed of 4 artificial 2-word phrases. Each artificial phrase was constructed by reversing the order of the 2 words in a real phrase.

We first evaluated ITPC spectrum for healthy individuals. In this study, the ITPC at each frequency was always normalized by subtracting the mean ITPC averaged over two neighboring frequencies. As shown in Fig. 2, significant response peaks were observed at the word rate (i.e., 2 Hz) and the phrase rate (i.e., 1 Hz) in both the real-phrase condition (2 Hz: P = 0.0009, false discovery rate (FDR)-corrected; 1 Hz: P = 0.0009, FDR-corrected) and the artificial-phrase condition (2 Hz: P = 0.0006, FDR-corrected; 1 Hz: P = 0.0006, FDR-corrected). Furthermore, significant responses were observed at frequencies that were harmonically related to the word and phrase rates. The harmonics were salient at 4 and 6 Hz, and remained clearly observable above 10 Hz. For EMCS patients, words-rate and phrase-rate responses were also significant in both the real (2 Hz: P = 0.0015, FDR-corrected; 1 Hz: P = 0.0266, FDR-corrected) and artificial phrase conditions (2 Hz: P = 0.0425, FDR-corrected; 1 Hz: P = 0.0266, FDR-corrected). Similarly, significant responses were observed at frequencies that were harmonically related to the word and phrase rates.

**Figure 2.**
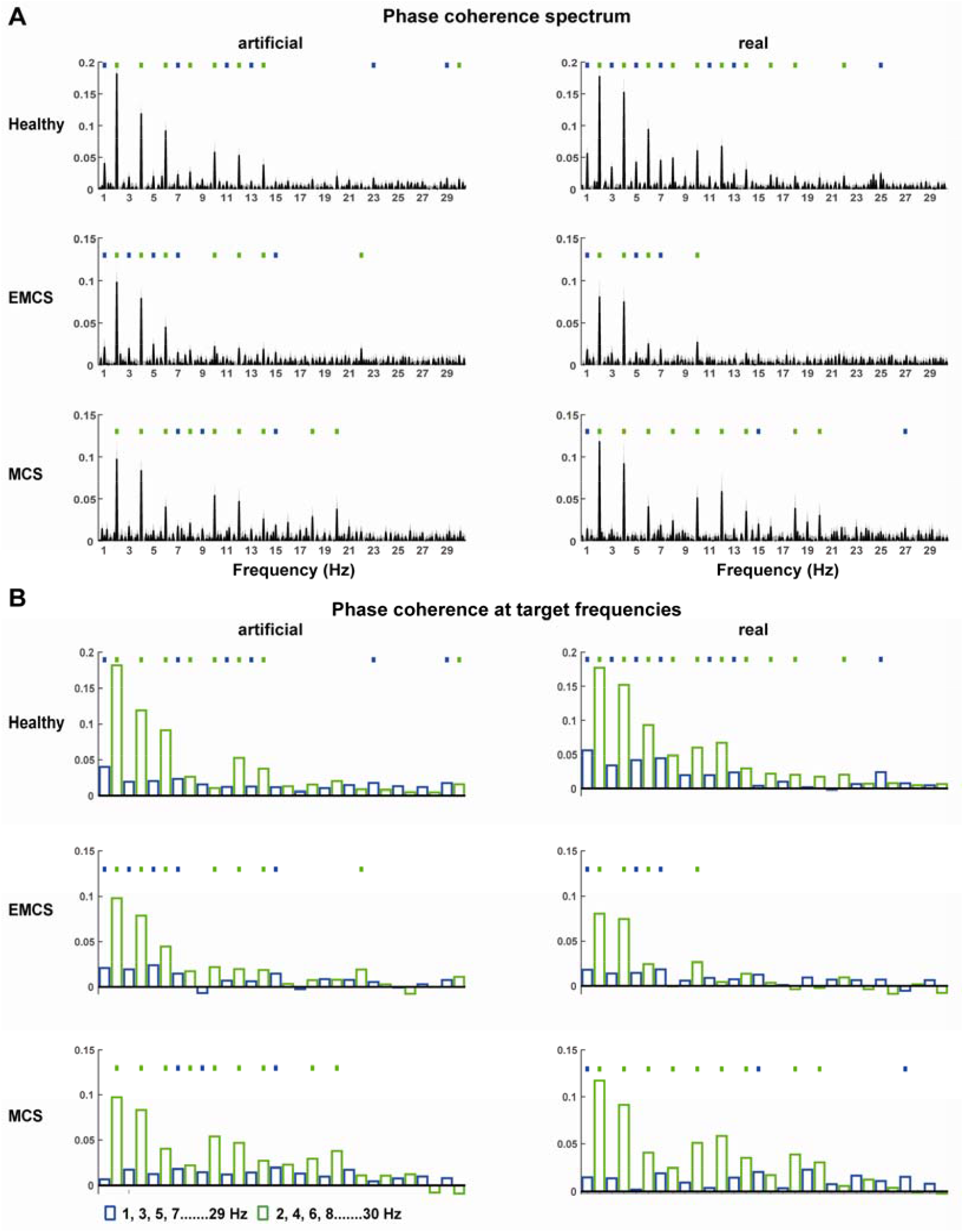
ITPC for each participant group. The ITPC at each frequency is normalized by subtracting the mean ITPC averaged over two neighboring frequencies. (A) The phase coherence spectrum for the artificial phrase condition (left) and real phrase condition (right). The spectrum was averaged over all EEG electrodes. The ITPC spectrum shows peaks at the word/phrase rate and harmonically related frequencies, which are further illustrated in panel B. (B) ITPC at target frequencies. If the ITPC is significantly higher than chance, it is marked by a dot.

For MCS patients, a significant phrase-rate response peak was observed in the real-phrase condition (1 Hz: P = 0.0224, FDR-corrected), but not in the artificial-phrase condition (P = 0.6498, FDR-corrected). Nevertheless, significant neural responses were observed at frequencies that were harmonically related to the word-rate in the artificial-phrase condition. The odd-order harmonics of the phrase-rate response, e.g., the 7-Hz, 9-Hz, and 15-Hz responses (P = 0.0240, 0.0240, and 0.0010, FDR-corrected, respectively), could not be explained by the harmonics of the word-rate response and therefore demonstrated that the brain could encode features related to the artificial phrases in the MCS state.

In the following, we separately analyzed the harmonics that could only be caused by the phrase-rate response, i.e., the responses at 3 Hz, 5 Hz, 7 Hz, etc., and the harmonics that could be caused by both the phrase- and word-rate responses, i.e., the responses at 2 Hz, 4 Hz, 6 Hz, etc. These two kinds of harmonics were separately referred to as *odd-order harmonics* and *even-order harmonics*. Furthermore, since there were a large number of harmonics, we grouped them into delta/theta-band harmonics (4-7 Hz), alpha-band harmonics (8-13 Hz), and beta-band harmonics (14-30 Hz). The harmonics falling into each band were averaged. As shown in Fig. 3, in the artificial-phrase condition, the mean ITPC of the odd-order harmonics in MCS patients was significant in all 3 bands (P = 0.0046, 0.0174, and 0.0174, for the theta, alpha, and beta bands, respectively FDR-corrected). In the real-phrase condition, the mean ITPC of odd-order harmonics in MCS patients was also significant in beta band (P = 0.0062, FDR-corrected).

**Figure 3.**
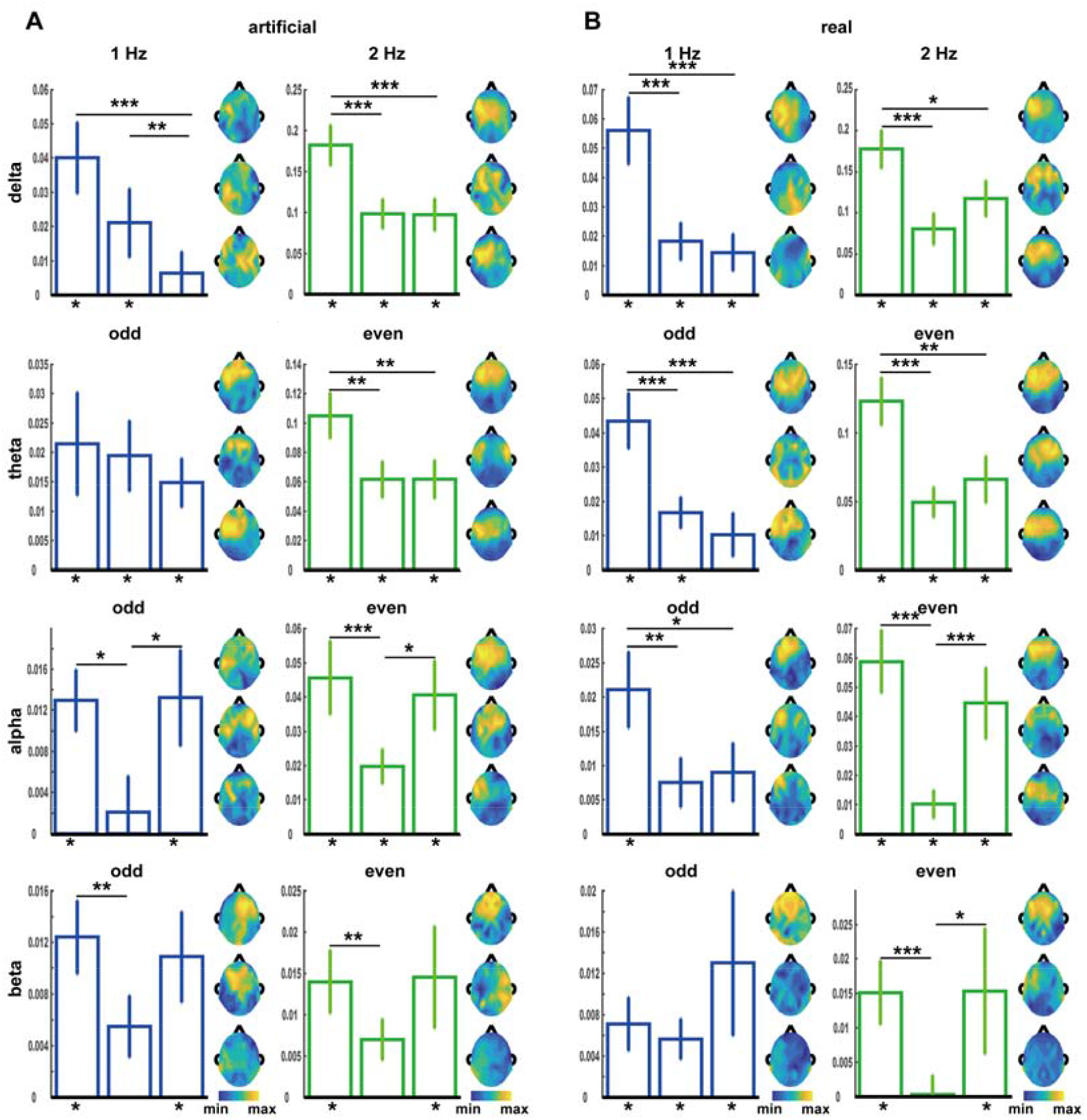
Comparisons of the ITPC across participant groups. The histogram from left to right and the response topography from top to bottom refer to the ITPC in healthy group, EMCS group, and MCS group in turn. The ITPC at the word and phrase rates are shown on the top row. The odd- and even-order harmonics are averaged into 3 frequency bands, i.e., theta (4-7 Hz), alpha (8-13 Hz), and beta (14-30 Hz) and shown in the 2^nd^-4^th^ row. Significant differences between populations were indicated by stars on top of each graph. (^*^P < 0.05, ^**^P < 0.01, ^***^P < 0.001, bootstrap, FDR corrected). The star underlying each bar indicates whether the ITPC is significantly higher than chance (P < 0.05, permutation test, FDR corrected). MCS, minimally conscious state; EMCS, emergence from the minimally conscious state.

We then analyzed whether the response ITPC differed across participant groups or conditions (Table. 1). A 2-way ANOVA (condition by group) analysis revealed that the phrase-rate ITPC significantly differed across groups but not between conditions. No significant interaction was observed between the two factors. Similar results were observed for the word-rate ITPC and the harmonic responses. For the phrase/word-rate responses and their theta-band harmonics, the ITPC was generally higher for healthy individuals than the other two populations. For the alpha- and beta-band harmonics, however, the even-order harmonics was stronger for MCS than EMCS patients.

**Table 1.**
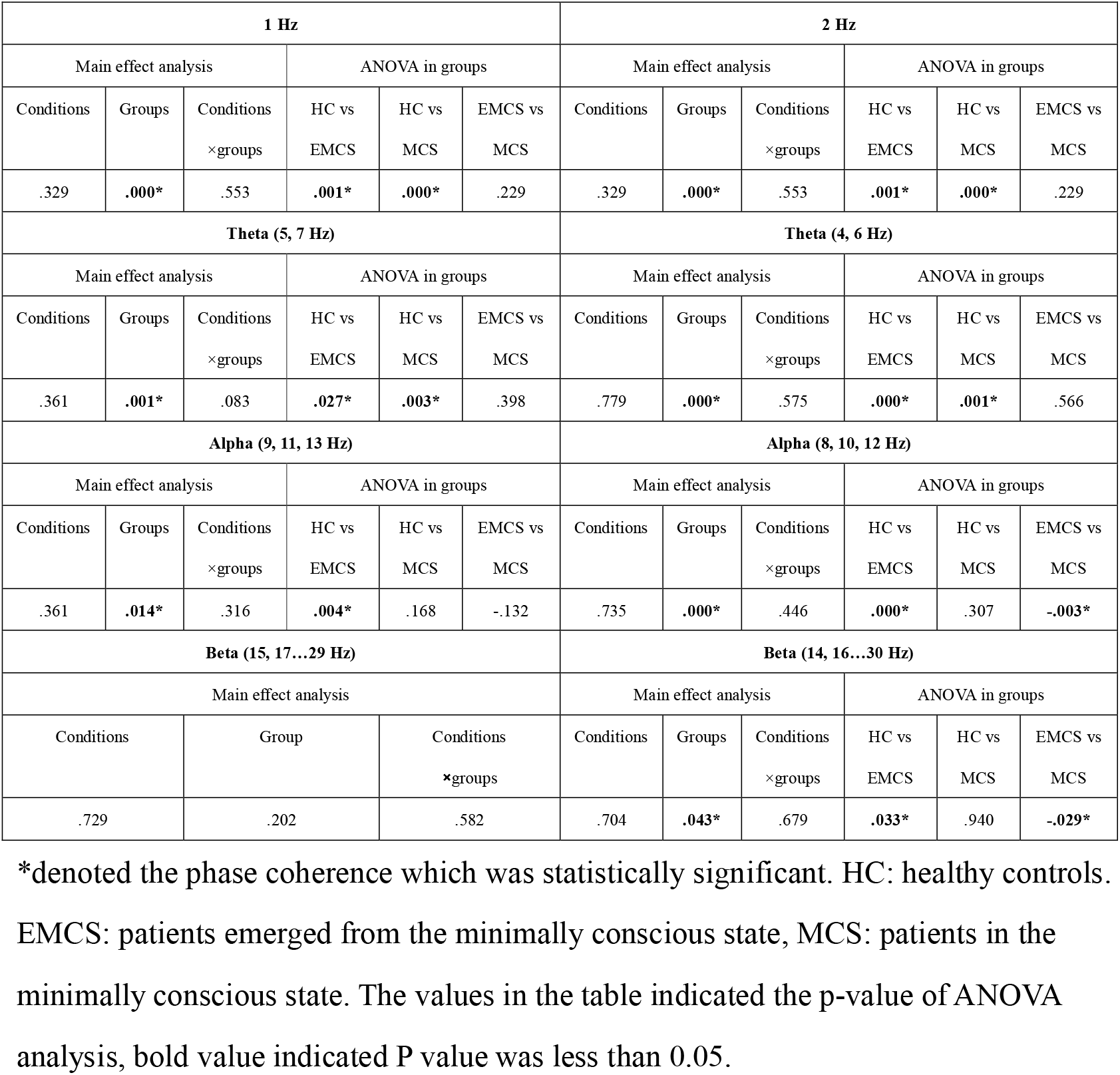
Two-way ANOVA analysis (condition by group) of ITPC.

## Discussion

Using a frequency-tagging approach, the current study demonstrated that statistical learning of artificial linguistic units can occur in a MCS state. Nevertheless, the neural representations of learned artificial phrases and real phrases were modulated by the consciousness state. Stimulus-locked neural responses in the delta and theta bands were enhanced by an increased level of consciousness. Stimulus-locked neural activity in the alpha and beta bands, however, showed a U-shaped curve, and is weakest in EMCS patients.

### Statistical learning in a MCS state

Statistical learning has been hypothesized as a fundamental mechanism for infants to learn the boundaries between words [1, 18]. It is established that, in the absence of prosodic cues related to word boundaries, infants could learn to segment a sequence of syllables into words [3, 4, 19]. For visual sequences, it is also shown that probabilistic regularities between items can be detected by both adults and infants [20-22]. Although SL is widely studied using behavioral and neural measures, a recent study has pointed out that most of these measures tend to be noisy and are only reliable at the group level [23]. Recent studies show that statistical learning engages widely distributed areas in cortex and also the hippocampus: The superior temporal gyrus primarily encodes transitional probabilities, inferior frontal gyrus and anterior temporal lobe primarily encodes ordinal position and identity, and the hippocampus primarily encodes identity [24]. Here, the transitional probability within each artificial phrase equaled to 1, and therefore the artificial phrases can also be referred to as artificial words. We referred to them as artificial phrases to be consistent to the common real phrases.

Previous studies suggest that, in healthy individuals, divided attention degrades but does not abolish the acquisition and consolidation of transitional probabilities [5, 25]. Furthermore, it is shown that sleeping neonates are able to automatically extract statistical properties of the speech input and thus detect the word boundaries in a continuous stream of syllables containing no morphological cues [26]. More recently, it is suggested that preverbal infants have the ability to track statistical regularities, infants and adults follow similar learning trajectories when tracking probability information among speech sounds [12]. These results indicate that although statistical learning can be modulated by attention it can occur largely automatically in healthy individual. The current study demonstrated that statistical learning can occur in patients in a MCS state, providing further evidence that statistical learning was a reliable process that occurs automatously. The phrasal response, however, was weaker in MCS patients than EMCS patients and healthy individuals, possibly reflecting disrupted frontal and parietal functions at reduced levels of awareness [27, 28].

### Low-frequency neural tracking of speech

When listening to speech, cortical activity can concurrently track speech units of different sizes, e.g., syllables, words, phrases, and sentences. It has been shown that even when the boundary between linguistic units is not associated with statistical cues, cortical activity can still track these linguistic units [7]. Nevertheless, when the linguistic units are purely defined by statistical cues, cortical activity tracking these linguistic units can also gradually emerge [9, 10]. Here, we presented both real phrases and artificial phrases that were purely defined by statistical cues, it was found that the phrasal-rate response to real and artificial phrases had comparable strength. This result suggested that when the number of phrases was limited (*N* = 4 in the current study) the phrasal-rate response was primarily driven by statistical cues. When the number of phrases was large so that the same phrase rarely repeat, the phrasal-rate response was more likely driven by other processes such as syntactic processing.

### Low-frequency neural activity and DoC

In the current study, the artificial phrases were not defined by prosodic features and could only be constructed by analyzing the statistical relationship between words. The real phrases could be parsed by either syntactic rules or by analyzing the statistical relationship between words. Therefore, neural tracking of artificial and real phrases cannot originate from neural encoding of acoustic features of speech but instead must reflect higher-level processes such as cortical encoding of statistical relationship between words or the neural construction of multi-word chunks. The result showed that low-frequency, i.e., 1-Hz, neural activity can track artificial and real phrases in healthy individuals and EMCS patients, but not in MCS patients.

This result was consistent with the hypothesis that slow cortical activity contributes to the emergence of consciousness [29] and attention [30], and was also consistent with previous results showing that MCS and UWS patients cannot track real spoken phrases [13, 15]. Furthermore, the results were also consistent with previous findings that low-frequency neural tracking of phrases is disrupted by sleep and lack of top-down attention [31, 32]. The formation of slow oscillations relies on the balance between neural excitation and inhibition [33, 34] and generally requires synchronization between large-scale neural networks [35, 36]. Slow cortical activity might be a neural substrate to integrate information across broad cortical areas and therefore may contribute to the conscious experience, which is always a unitary and undivided whole [37, 38].

Here, an increase in delta- and theta-band activity is associated with an increase in consciousness level. In contrast, some previous studies have reported an increase in slow oscillations in anesthetized participants [39] or unresponsive participants [40, 41]. The slow oscillations in these studies, however, are generally spontaneous, non-stimulus-locked oscillations, instead of the stimulus-locked response analyzed in the current study. Therefore, it is possible that the ability to synchronize low-frequency neural activity to external stimulus, instead of the absolute power of low-frequency activity, is an index of consciousness level.

Prolonged disorders of consciousness (DoC) are defined as any disorder of consciousness that has continued for at least four weeks following sudden onset brain injury, includes patients in coma, VS/UWS, and MCS [42, 43]. It is established that high level, covert cognition may be present in patients whose bedside evaluation appears consistent with the vegetative state or minimally conscious state [44-46]. Identification of covert cognition in patients with prolonged DoC is important, as patients with covert consciousness (cognitive motor dissociation, CMD) have a better prognosis than other patients, determining residual cognition in these patients has its crucial impact on family counselling, decision-making and the design of rehabilitation programs [47-50]. Therefore, it is important to generalize diagnostic tools to identify CMD patients for patients with prolonged DoC. Currently, the covert consciousness is typically probed with paradigms that require the patient to follow repeated commands to imagine that they are moving. The high demands on cognitive ability result in low sensitivity in the aspect of detecting CMD [50]. In this study, we found that statistical learning abilities were preserved in consciousness perturbation, indirectly indicating that the EEG response during passive statistical learning may be a promising paradigm to detect residual high-level cognition in patients with DoC.

In conclusion, we found that statistical learning abilities was preserved in consciousness perturbation. Furthermore, the study shows that neural activity tracking spoken words and phrases showed a large number of harmonics in the theta, alpha, and beta bands. An increase in delta- and theta-band stimulus-locked neural activity and a decrease in alpha- and beta-band stimulus-locked neural activity are suggestive of an improvement of consciousness in DoC patients.

## Data Availability

All data produced in the present study are available upon reasonable request to the authors

## Acknowledgements

The authors would like to thank the patients, their families, and volunteers for their participation in this study. This work was supported by National Natural Science Foundation of China 81870817, Guangzhou Key R&D Program of China 202007030005, National Key R&D Program of China 2018YFA0701400, and supported by the MOE Frontier Science Center for Brain Science & Brain-Machine Integration, Zhejiang University.

